# Exploration of cortical ß-Amyloid load in Alzheimer’s disease using quantitative susceptibility mapping at 9.4T

**DOI:** 10.1101/2022.09.23.22280290

**Authors:** Elisa Tuzzi, Alexander Loktyushin, Anja Zeller, Rolf Pohmann, Christoph Laske, Klaus Scheffler, Gisela E. Hagberg

## Abstract

Early detection of β-Amyloid (Aβ) deposits in Alzheimer’s disease (AD) patients may enable treatment in the early stages of the disease. To date, there are no validated, specific, and non-invasive routines for early Aβ detection which are suitable for clinical practice. Ultra-high resolution quantitative susceptibility mapping (QSM) at 14.1T has previously shown different contrasts in cortical areas of an AD sample that resembled distinct Aβ spatial patterns in histological sections of the same specimen. These contrasts appeared different in the QSM from a healthy control (HC) sample where, instead, no plaques were detected. In a few cases, this distinction in the cortical magnetic effects (para- and diamagnetic) between AD and HC was also observed in vivo using ultra-high resolution single-echo Acquisition Weighted (AW) gradient echo MRI at 9.4T. Based on this evidence, a method to quantify the paramagnetic and diamagnetic effects of Aβ to possibly distinguish between AD and HC was developed. In this study, we extended those results and explored the ability to use QSM to estimate β-Amyloid plaque load in the cortex of 7 elderly patients with early AD and 7 healthy age-matched HC. Besides ultra-high resolution AW images, we acquired lower resolution multi-echo (MTE) data and compared the previously used RESHARP- based method for background removal (“AW-filter”, with a high Tikhonov term and a small kernel size) with a method widely used in the literature (“mild-filter”, with a low-regularization term and a large kernel size) for the MTE processing. Paramagnetic and diamagnetic QSM changes were assessed in 16 cortical areas. All methods enabled the detection of regions with high QSM values (up to 45 ppb in AD and up to 25 ppb in HC) and known as early Aβ accumulation areas in AD progression. A distinct cortical pattern was observed at both spatial resolutions using the AW-filter. This was not the case with the mild-filter at the lower resolution. AW-QSM outperformed MTE maps with the AW-filter for the detection of areas with prominent cortical paramagnetic effects, including regions where Aβ accumulation happens in the earliest AD stages, such as the precuneus and posterior cingulate cortex. Diamagnetic changes were more prominent than the paramagnetic effects regardless of the spatial resolution used and this difference was further enhanced with the mild-filter. This explorative study points toward the development of more accessible clinical methods to non-invasively detect effects of Aβ accumulation in AD patients by exploring cortical features that can be detected by ultra-high field QSM at different spatial resolutions.

## 1. INTRODUCTION

Alzheimer’s disease (AD) is a progressive brain pathology currently affecting over 50 million people worldwide and is the most common cause of dementia. AD is characterized by an abnormal accumulation of β-Amyloid (Aβ) deposits (or plaques) in the brain tissue starting several years before the clinical onset [1–6], with diffuse and fibrillar types prevailing in the preclinical and end-stages, respectively [7]. According to Braak neuropathological stages [8], Aβ deposits begin to aggregate in the basal portion of the isocortex and then spread out through the association areas to finally include the primary isocortical areas at the end-stage. The more recent Thal amyloid phase [9], describes β-Amyloid accumulation starting from the neocortex (phase I), developing to the allocortex in the phase II (hippocampus, entorhinal cortex and cingulate gyrus), and including the basal ganglia (phase III), midbrain (phase IV) and cerebellum (phase V) in the later stages. The detection of Aβ plaques in its very early stage may enable possible treatment in the presymptomatic phase thus preventing the onset of the disease. Therefore, it is crucial to identify routines for early Aβ detection which should ideally be specific, non-invasive and suitable for the clinical practice. Although several potential biomarkers for AD pathology have been evaluated [10, 11], to date, there are no validated specific and non-invasive clinical routines for early Aβ detection. The current established techniques for Aβ plaque load detection in vivo are Aβ-Positron Emission Tomography (PET) imaging with Pittsburgh compound-B [^11^C]-PIB, [^18^F] florbetapir, [^18^F] flutemetamol and [^18^F] FC119S tracers in combination with Aβ_1-42_ Cerebrospinal fluid (CSF) [12–13] quantification. However, PET imaging involves exposure to ionizing radiation besides the fact that tracers bind non-specifically to fibrillar, dense-cored, and diffuse plaques [14]. Furthermore, Aβ_1-42_ CSF test is not a well-accepted practice yet [15] and lacks standardized protocols across assays [16]. On the other hand, quantitative magnetic resonance imaging (MRI) at ultra-high magnetic field strengths (UHF), with the currently increasing availability of 7T scanners in the clinical practice, is an emerging non-invasive tool for the assessment of pathological microstructural alterations in brain tissue by enabling very high resolution imaging. UHF-MRI may therefore provide a potential non-invasive means for distinguishing AD from HC in the early stages of Aβ accumulation by detecting cortical changes in AD patients directly linked to the plaques. Indeed, β-Amyloid proteins, which manifest diamagnetic properties [17] and likely induce alterations in the myelin structure (diamagnetic), can bind to iron (paramagnetic) causing local in-homogeneities in the static magnetic B_0_ field. These changes can be detected in the phase images acquired with gradient-echo (GRE) sequences. The spatial distribution of the tissue magnetic susceptibility (χ) can then be derived from the measured magnetic field perturbations using post-processing technique for retrieving quantitative susceptibility maps (QSM). Studies showing the use of QSM to characterize Alzheimer’s disease are emerging [18]. Although most of these studies reported increased QSM values in subcortical structures, cortical alterations have also been reported at 3 and 7T [19–23]. Ultra-high resolution QSM at 14.1T showed different magnetic bands across the cortex of an AD and a HC specimen that reflected the spatial patterns in Anti-Aβ Immunohistochemistry of the same samples [24]. In the same study, distinct cortical patterns were also observed in vivo in 2 AD patients and 2 HC subjects at 9.4T. Based on those observations, a method to quantify paramagnetic and diamagnetic effects across the cortex of AD patients was developed to investigate the possibility of distinguishing AD from HC based on ultra-high resolution QSM.

In the present study, our first aim was to confirm those results to a larger cohort of elderly patients with early AD and age-matched HC. Our second aim was to explore the ability to detect the same effects using lower resolution data. We therefore explored and extended our previous observational study from QSM obtained with voxel-sizes close to the plaque size at 14.1T to larger voxel-sizes achievable in vivo at 9.4T. These steps are aimed at exploring tissue features that can be detected in AD patients using QSM at different spatial resolutions and magnetic field strengths. Our results can potentially open up future possibilities for the clinical translation of QSM methods that aim at detecting cortical alterations of the pathological tissue affected by Aβ at lower magnetic field strengths.

## 2. METHODS

QSM images were generated from an ultra-high resolution single-echo GRE acquisition weighted (AW) sequence [25] (as previously used [24]) in 7 elderly patients with early AD and 7 matched HC, and from a lower spatial resolution multi-echo (MTE) sequence, with two strategies for background (BG) removal based on the RESHARP algorithm [26] prior to QSM calculation. Specifically, we first employed the same filter as we used in our previous study, with a high regularization term and a small spatial kernel, namely “AW-filter”, and then a “mild-filter”, with a low regularization term and a large kernel size. In total, we therefore explored three methods for the evaluation of the paramagnetic and diamagnetic effects in the cortex of AD patients. The paramagnetic load was assessed after exclusion of susceptibility values > 50 ppb likely coming from the veins [27]. Paramagnetic and diamagnetic effects were assessed across the cortex and in specific cortical regions of interest (ROIs) obtained from the Harvard-Oxford Cortical Atlas [28].

### 2.1. Subjects

We examined seven patients with early AD (5 Female, 2 Male; 74 ± 8.06 years) and 7 HC (3 Female, 4 Male; 72.00 ± 8.69 years) in a 9.4T whole-body human-research MRI system (Siemens Healthineers, Erlangen, Germany), equipped with a custom-built RF-coil-array (16 transmit channels and 31 receive channels) optimized for homogeneity and signal-to-noise ratio inside the human head [29]. The measurement protocol included three optimized acquisition sequences for a total duration of 32min and 54s. The study was approved by the Ethics Review Board of the Eberhard Karl’s University of Tübingen and all subjects gave their written informed consent to participate. AD patients were diagnosed based on an array of neuropsychological and clinical tests including CSF total tau, CSF phosphorylated tau and CSF Aβ. Other tests included: BDI (Beck Depression Inventory), GDS (Global Deterioration Scale), MMSE (Mini Mental State Examination), CEDAR (Consortium to Establish a Registry for AD), which were also administered to the healthy controls.

### 2.2. MRI acquisition protocol

A whole brain Magnetization Prepared 2 Rapid Gradient Echo (MP2RAGE) contrast was used as anatomical reference in the native space of each subject: inversion times = TI1/TI2 = 900/3500 ms; echo time (TE) = 2.3 ms, flip angle (α°) = 4°/6°; read-out repetition time (TR) = 6 ms; voxel-size = (800 µm)^3^, matrix size = 256x256x192, field of view (FOV) = 205x205x154 mm^3^, acquisition time (TA) = 8 min and 53 s. These images were then corrected for deviations of the transmit field [29]. Two different sequences were acquired for QSM calculation:

1. A partial-coverage, small slab thickness, flow-compensated, ultra-high resolution single-echo 3D gradient-echo (GRE) Acquisition Weighted (AW) sequence with weighted averaging of phase-encoding steps [25] (voxel-size = 132x132x.610 μm^3^, FOV = 180x135x12.8 mm^3^, reconstructed matrix = 1362x1024x21, TR/TE/α° = 24/16.5 ms/8°, TA = 14.75 min). The 12.8 mm thick slab was carefully placed to include the frontal, parietal, temporal and occipital cortices as well as the basal ganglia, representing early Aβ accumulation areas [8, 9, 31].
2. A multi-echo (MTE) 3D-GRE with FOV = 168x192x77 mm^3^, voxel-size = 375x375x800 μm^3^, reconstruction matrix = 448x512x96, GRAPPA factor of 2 in the first phase-encoding dimension, elliptic trajectory and partial Fourier coverage of 6/8 in the phase-encoding dimensions, TR = 41 ms, TEs = 5.93/12.62/17.92/23.22/28.52/33.82 ms, α◦ = 11◦ and TA = 8.5 min. Additional navigator signals before and after acquisition of the gradient echoes were used to detect and correct variations in the signal phase due to fluctuations of the magnetic field. 3D image reconstruction including the navigators was obtained as described in [32, 33]. The FOV orientation was the same as for the AW image.

### 2.3. AW-based QSM reconstruction

Quantitative susceptibility maps from the ultra-high resolution AW sequence were obtained using the same pipeline as previously described [24]. Specifically, first, complex images were computed using an optimized method for coil sensitivity evaluation and adaptive combination of the channels [34]. A Laplacian-based unwrapping algorithm, as implemented in the MEDI-toolbox (http://pre.weill.cornell.edu/mri/pages/qsm.html) was used to correct the phase wraps of the unmasked phase images [35]. For background field correction, a brain mask was created using the brain extraction tool (BET) as implemented in FSL (http://www.fmrib.ox.ac.uk/fsl/) [36] with a threshold for the fraction intensity ranging between 0.01 and 0.05, depending on the subject. The whole brain was used as a reference region for QSM calculation according to the literature [18, 37] by subtracting the median value of the unwrapped phase images inside the brain mask from all the unwrapped phase image values. Background field removal of the masked unwrapped phase images after median subtraction was achieved using the RESHARP algorithm [26] with a Tikhonov regularization parameter of 10^-3^ and a kernel size set to 2 times the voxel-size in the second phase-encode dimension (1.22 mm). The iterative least squares (iLSQR) approach was then applied for the QSM computation, as implemented in the STI-Suite v2.2 [38], using 15 iterations and 40% zero-padding in the second phase encode dimension.

### 2.4. MTE-based QSM reconstruction

Susceptibility maps from the lower resolution MTE GRE sequence were reconstructed using the same workflow as the AW-GRE sequence, with slight modifications. Complex images were computed using an ASPIRE-like algorithm [39]. Single-echo phase data were used for QSM calculation to generate more accurate and homogeneous susceptibility maps [40]. Specifically, phase data from the third echo (17.92 ms) was chosen as it provided the highest grey/white matter contrast in the QSM images compared to combinations of the echoes and corresponds to the TE with the highest signal-to-noise ratio (SNR) based on our previously reported R_2_* values of 60-70s^-1^ in the AD patients’ cortex [24]. To correct for the wraps of the unmasked phase images a Laplacian-based unwrapping algorithm, as implemented in the STI toolbox (http://people.duke.edu/~cl160/), was used as for the AW-data. The RESHARP algorithm [26] was used with two sets of optimization parameters: a high Tikhonov regularization term of 10^-3^ with a small kernel size of 1.22 mm, as for the AW-data, and a small Tikhonov parameter of 10^-12^ in combination with a large kernel size of 1.6 mm (2 times the voxel-size in the second phase-encode dimension), which maximizes iron-dependent QSM contrast using lower voxel-sizes at 9.4T, as previously described [33].

### 2.5. Registration of the Harvard-Oxford Cortical ROIs to the AW native space

In order to extract the susceptibility values in the AW native space of each subject, the 48 bilateral FSL-Harvard Cortical ROIs in MNI-space were spatially registered to the ultra-high resolution AW space of each subject, through the following steps:

1. First, the B_1_-corrected MP2RAGE contrast image was spatially normalized to MNI-space using DARTEL in SPM12 (http://www.fil.ion.ucl.ac.uk/spm/), as previously described [30].
2. The inverse DARTEL transformation was used to bring the 48 bilateral FSL Harvard-Oxford ROIs into the native space of the MP2RAGE of each subject.
3. The AW images were registered to the native space of MP2RAGE images of each subject through a 4-step process (see Section 2.6., below).
4. ROIs in the native MP2RAGE space of each subject were re-sliced to the AW images obtained in step 3) using SPM12 (http://www.fil.ion.ucl.ac.uk/spm/).
5. Finally, by inverting the process used in step 3) the ROIs in the MP2RAGE space were registered to the AW native space of each subject.

### 2.6. Registration of the AW native space to the MP2RAGE native space

To optimize the spatial registration between the small slab thickness (12.8 mm) AW image and the whole brain MP2RAGE contrast, we performed the following steps:

1. Spatial reorientation and registration of the bigger FOV magnitude image from the third echo of the MTE sequence to the first inversion time image of the MP2RAGE sequence using, respectively, SPM12 (http://www.fil.ion.ucl.ac.uk/spm/) and a rigid-body transformation as implemented in the Advanced Normalization Tools (ANTs) (https://sourceforge.net/projects/advants/files/) [41].
2. Spatial reorientation and registration of the AW magnitude image to the third echo MTE image as obtained in step 1) using, respectively, SPM12 (http://www.fil.ion.ucl.ac.uk/spm/) and the same rigid-body ANTs-based transformation as used in step 1).
3. The AW image obtained in step 2) was then registered to the MP2RAGE space by applying the transformation obtained in step 1) for the spatial registration of the MTE magnitude image to the MP2RAGE space.

### 2.7. Definition of the Cortical ROIs in AW image space

To compare the results obtained with all methods at both spatial resolutions within the same ROIs in all subjects, we first spatially registered the ROIs in the AW space to the ROIs in the MTE space using ANTs. Then, due to the reduced size of the AW-FOV (180x135x12.8mm^3^) which did not include all the 48 bilateral cortical Harvard ROIs and the misalignment across subjects, common ROIs had to be defined in the AW space in a first place. The same subset of ROIs was then used for the analyses in both AW and MTE native spaces.

The following steps describe the process to obtain the cortical ROIs which were common to all subjects in the AW space.

1. First, the tissue probability maps of grey and white matter (GM, WM) as well as the cerebrospinal fluid (CSF) segmented from quantitative T_1_-maps (obtained from MP2RAGE images) using SPM12 (http://www.fil.ion.ucl.ac.uk/spm/) were registered to the AW native space using the same process as described in **2.6.4.** and **2.6.5.** for the cortical ROIs.
2. The GM, WM and CSF tissue probability maps in the AW space were summed to obtain a preliminary brain mask.
3. A final GM brain mask was obtained by multiplying the mask obtained in step 2) by the GM tissue probability map thresholded at 0.9 to achieve a more conservative mask and thus reducing partial volume effects.
4. The cortical ROIs in the AW space were multiplied by the final GM brain mask obtained in step
5. 3) to create the final “ROI binary masks” which included only voxels located inside the GM.
6. QSM images obtained after excluding susceptibility values higher than 50 ppb, attributed to the veins [27], were multiplied by the ROI binary masks.
7. All the voxels with a non-zero susceptibility value within each ROI binary mask were summed up to calculate the “nvoxROI”, for each ROI.

Finally, a ROI was considered common to all subjects in the AW native space when its number of voxels with non-zero susceptibility values, i.e. “nvoxROI”, differed from zero. Sixteen out of 48 bilateral Harvard-Oxford Cortical ROIs showed nvoxROI≠0 in all subjects and were therefore used for the analyses: the Frontal Pole, the Inferior Temporal Gyrus temporo-occipital part, the Inferior Frontal Gyrus pars opercularis, the Precentral Gyrus, the Postcentral Gyrus, the Supramarginal Gyrus anterior division, the Supramarginal Gyrus posterior division, the Angular Gyrus, the Lateral Occipital Cortex superior division, the Paracingulate Gyrus, the Cingulate Gyrus anterior division, the Cingulate Gyrus posterior division, the Precuneous Cortex, the Cuneal Cortex, the Frontal Operculum Cortex and Central Operculum Cortex.

### 2.8. Registration of the Harvard-Oxford Cortical ROIs to MTE native space

The same subset of ROIs defined in the AW space was used for the analyses in the MTE spaces. To this purpose, the FSL-Harvard Cortical ROIs in the MP2RAGE native space of each subject (as obtained in **2.5.2.**) were registered to the MTE native space by applying the inverse ANTs-based rigid transformation used to register the MTE to the MP2RAGE native spaces.

### 2.9. Vein fraction calculation

In both the AW and MTE spaces, a “vein mask” inside each ROI was created by thresholding QSM at 50 ppb (χ>50 ppb) and then multiplying the result by the ROI binary mask. The “vein fraction” inside each ROI was then defined as the number of pixels of the vein mask divided by the number of pixels of the ROI binary mask. In a similar way, the vein fraction inside the cortex (only including the 16 common ROIs) was also calculated. A two-sample t-test was computed to test significant differences in the vein fraction between AD and HC groups for each method.

### 2.10. Paramagnetic load calculation in the AW and MTE native spaces and statistical tests

Sixteen common cortical ROIs were defined in the AW and MTE native space (see section **2.7.**) of each subject. The ratio of the number of paramagnetic pixels (χ>0) inside each ROI to the total number of pixels of the ROI binary mask was then computed for several QSM cutoff-values, ranging from 10 to 50 ppb. This approach yielded a curve describing the paramagnetic pixels as a function of QSM cutoff-values (paramagnetic load), as previously described [24]. The paramagnetic load was calculated for QSM obtained at both spatial resolutions (**2.3.** and **2.4.** sections) after exclusion of the vein contribution (χ>50 ppb) [27]. The lower threshold of 10 ppb was set based on our previous results showing this cutoff-value correctly identifying a paramagnetic plaque-fraction of 0.15 (15% of the cortical considered area) in the corresponding histological section of an AD specimen [24]. For each subject, the paramagnetic load across the cortex (containing the 16 ROIs) was also calculated using the same approach. A two-sample t-test (α=0.05) was used to test significant differences between the mean paramagnetic load of the two groups (AD and HC) in each cortical ROI and in the cortex. Bonferroni correction for multiple comparisons of both para- and diamagnetic effects within the 16 ROIs was also applied (α=0.05/32). The same approach was used to estimate the paramagnetic load from the ROIs in the MTE space.

### 2.11. Diamagnetic effects calculation in the AW and MTE native spaces and statistical tests

Similarly to the paramagnetic load, the number of diamagnetic pixels (χ<0) inside each ROI was divided by the number of pixels of the ROI binary mask for several QSM cutoff-values (from -50 to - 10 ppb) to obtain a curve describing the fraction of diamagnetic pixels as a function of the QSM cutoff-values. The same approach was used to evaluate the diamagnetic contribution inside the cortex. The diamagnetic effects were then averaged across the subjects of each group and a two-sample t-test was computed to test significant differences between AD and HC, with and without Bonferroni correction for multiple comparisons.

### 2.12. Histogram analysis

Normalized histograms were computed from the QSM images obtained using the AW and the MTE sequences inside the cortex and in the 16 cortical common ROIs, using the MATLAB function “histogram”. Significant differences between the AD and HC histogram curves were then tested using a two-sample Kolmogorov-Smirnov test, as implemented in MATLAB (“kstest2” function).

## 3. RESULTS

QSM obtained using both the AW and the MTE sequences are shown in Figure 1 for an AD patient (a, b, c) and a HC (g, h, i). The vein masks (χ>50 ppb) inside the cortex of the two subjects are also shown for the AW (d, l) and the MTE maps (e, m, f, n), respectively. In general, an enhanced contrast between grey and white matter (GM, WM) was observed in the QSM of the AD patients (with χ values up to 45 ppb) compared to HC (χ values up to 25 ppb), independently of the sequence and filter used. However, the AW-filter showed a distinct cortical pattern in the QSM images of the AD patients (Fig. 1, a, b), which was less evident with the mild-filter (Fig. 1, c). In contrast, the mild-filter in the MTE maps showed a strong contrast in subcortical regions which was almost lost at higher resolution. Furthermore, the spatial pattern of the voxels with high QSM values appeared sharper and thus more localized in the AW maps compared to the MTE maps. The contribution from veins (χ>50 ppb) in the cortex of the AD patient (Fig. 1, d, e, f) and the HC (Fig. 1, l, m, n) was almost absent in the AW maps compared to the lower resolution maps, where veins appeared smoothed and more extended with the mild than with the AW-filter. No significant difference was observed in the vein fraction between AD and HC. Figure 2 shows a representative ROI (the “*Supramarginal Gyrus posterior division*”, blue) overlaid on the QSM images of an AD patient (A, B, C) and a HC (G, H, I) obtained, respectively, with the AW (A, G) and the MTE sequences using the AW-filter (B, H) and the mild-filter (C, I). The spatial patterns of the paramagnetic effects obtained for χ>10 ppb (D, E, F, L, M, N, red boxes) and the diamagnetic contribution (χ<0 ppb; a, b, c, d, e, f) inside the cortex of the two subjects are also shown. Both effects (paramagnetic and diamagnetic) exhibited slight different spatial distributions depending on the voxel-size and the filter used. In general, the AW maps showed small-sized and localized effects compared to the spatially more extended effects detected at lower resolution. No significant difference was found between AD and HC when comparing the paramagnetic load across the cortex. In contrast, a significant difference (corrected and uncorrected p values) was found at the ROI-level when comparing the paramagnetic load of the AD patients with controls. Figure 3 shows the mean paramagnetic load curves for the AD (red) and HC (blue) groups obtained from the QSM at both resolutions, using the three methods (AW map (a), MTE map with AW-filter (b) and MTE map with mild-filter (c)). In the high resolution maps, eight out of sixteen cortical ROIs showed a significant difference between AD and HC groups based on the paramagnetic load (Table 1): the *Inferior Frontal Gyrus pars opercularis,* the *Supramarginal Gyrus anterior and posterior divisions,* the *Paracingulate Gyrus,* the *Cingulate Gyrus anterior division,* the *Cingulate Gyrus posterior division,* the *Precuneous Cortex,* and the *Frontal Operculum Cortex.* After correction for multiple comparisons, three of these regions still differed between groups: *the Paracingulate Gyrus, the Cingulate Gyrus posterior division, and the Frontal Operculum Cortex.* Four of these regions were also detected by the MTE maps: the *Inferior Frontal Gyrus pars opercularis,* the *Supramarginal Gyrus anterior* and *posterior divisions,* and the *Frontal Operculum Cortex*, and after correction only the *frontal operculum cortex* still exhibited a difference between groups. On the other hand, two regions were only detected in the MTE maps. As to the diamagnetic effects, six ROIs were detected regardless of the spatial resolution, while six additional regions were only detected with the MTE methods and two regions only with the AW technique. Overall, a consistent higher significance (p<<0.05/32) between groups was observed for the diamagnetic effects than the paramagnetic load, at both resolutions. In contrast, only one region showed a very high significant difference (p<<0.05/32) between the groups based on the paramagnetic effects in the AW maps. Two regions known to be involved in the earliest stage of Aβ accumulation, the precuneus and the posterior cingulate cortex, were only captured by the AW technique. Figure 4 shows the regions with a significant difference between AD and HC groups based on the paramagnetic load (a) and the diamagnetic effects (b) obtained with and without correction for multiple comparisons (“pcorr” and “puncorr”, respectively). The ROIs are superimposed on the same magnitude image of the AW sequence from an AD patient. Blue refers to the ROIs which were detected by both AW- and MTE-QSM regardless of the QSM effects (paramagnetic (a) or diamagnetic (b)). Specifically, four ROIs were detected for paramagnetic effects and six ROIs for diamagnetic changes. Red and yellow refer to the regions detected by only the AW technique and only the MTE maps, respectively. In total, ten regions were detected based on the paramagnetic load, while fourteen regions were captured depending on the diamagnetic effects. Results are resumed in Table 1. Histograms of the susceptibility values obtained from the QSM at both resolutions showed statistically significant differences (p<0.05) between AD patients and HC groups based on a two-sample Kolmogorov-Smirnov test, in five out of sixteen ROIs (Fig. 5). In general, broader curves with lower peaks were observed using the mild-filter in comparison to the AW-filter at lower resolution. Also, higher paramagnetic and diamagnetic values were always observed in the AD patients (red) compared to HC (blue) when using the mild-filter with respect to the other methods. In addition, a consistent slight shift towards higher paramagnetic effects was observed in the patient’s curve at lower resolution (more pronounced with the mild-filter) compared to the ultra-high resolution. Overall, smaller absolute values of the susceptibility were always prevalent in HC, regardless of the method, while higher absolute values generally prevailed in patients.

**Fig. 1.**
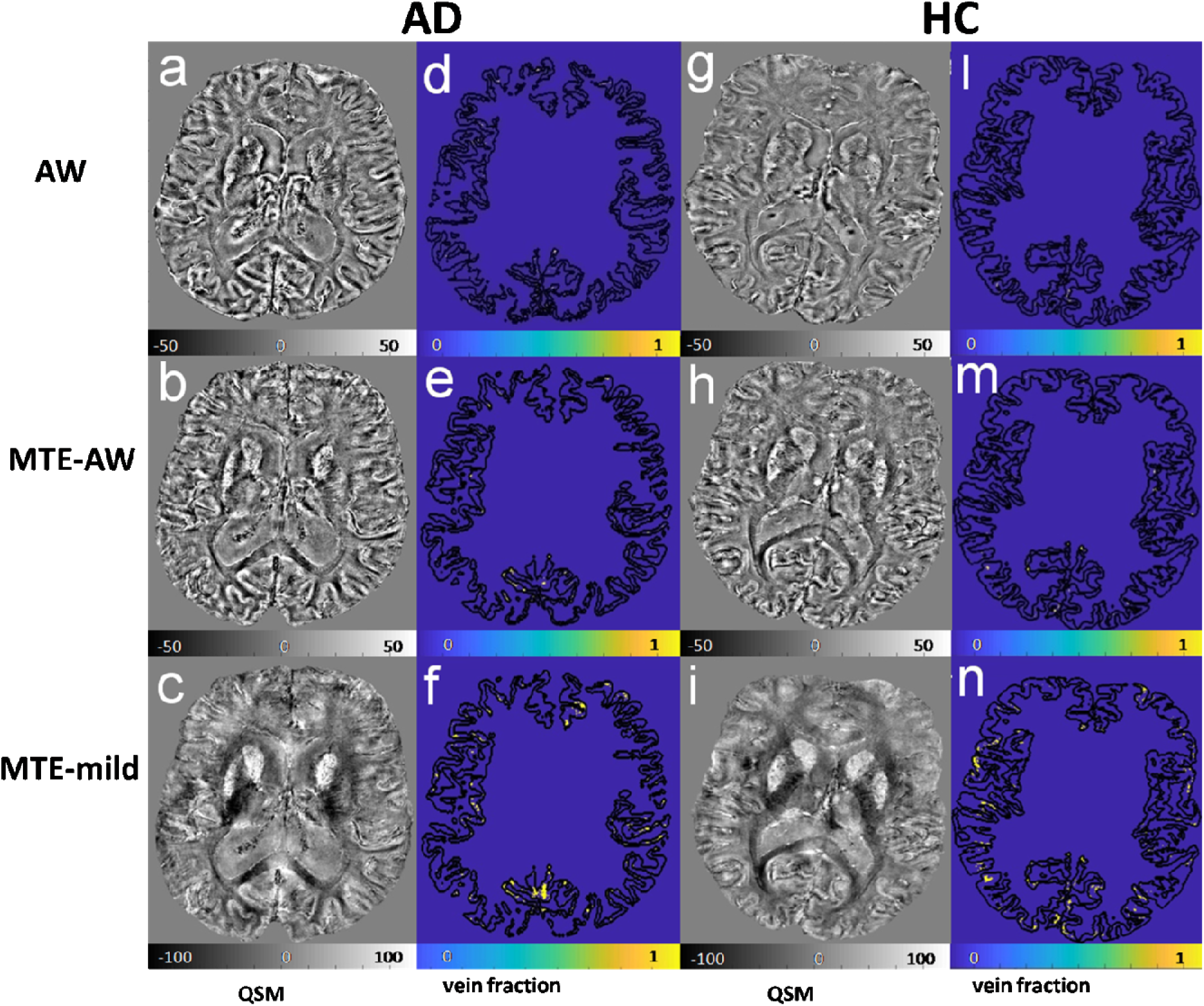
QSM (a, b, c, g, h, i) and cortical vein masks (χ>50 ppb; d, e, f, l, m, n) of an AD patient (a-c, d-f) and a HC (g-i, l-n) obtained from ultra-high resolution (“AW”, a, d, g, l) and lower resolution QSM using the AW-filter (“MTE-AW”, b, e, h, m) and the mild-filter (“MTE-mild”, c, f, i, n).The AW-filter yielded a distinct cortical pattern with increased QSM values in cortical regions (see zoomed image in Fig. 2) but lower susceptibility values than the mild-filter in the lentiform nucleus. **Abbreviations: AW**, ultra-high resolution sequence; **MTE-AW**, map obtained from the MTE sequence and using the AW-filter; **MTE-mild**, map obtained from the MTE sequence and using the mild-filter.

**Fig. 2.**
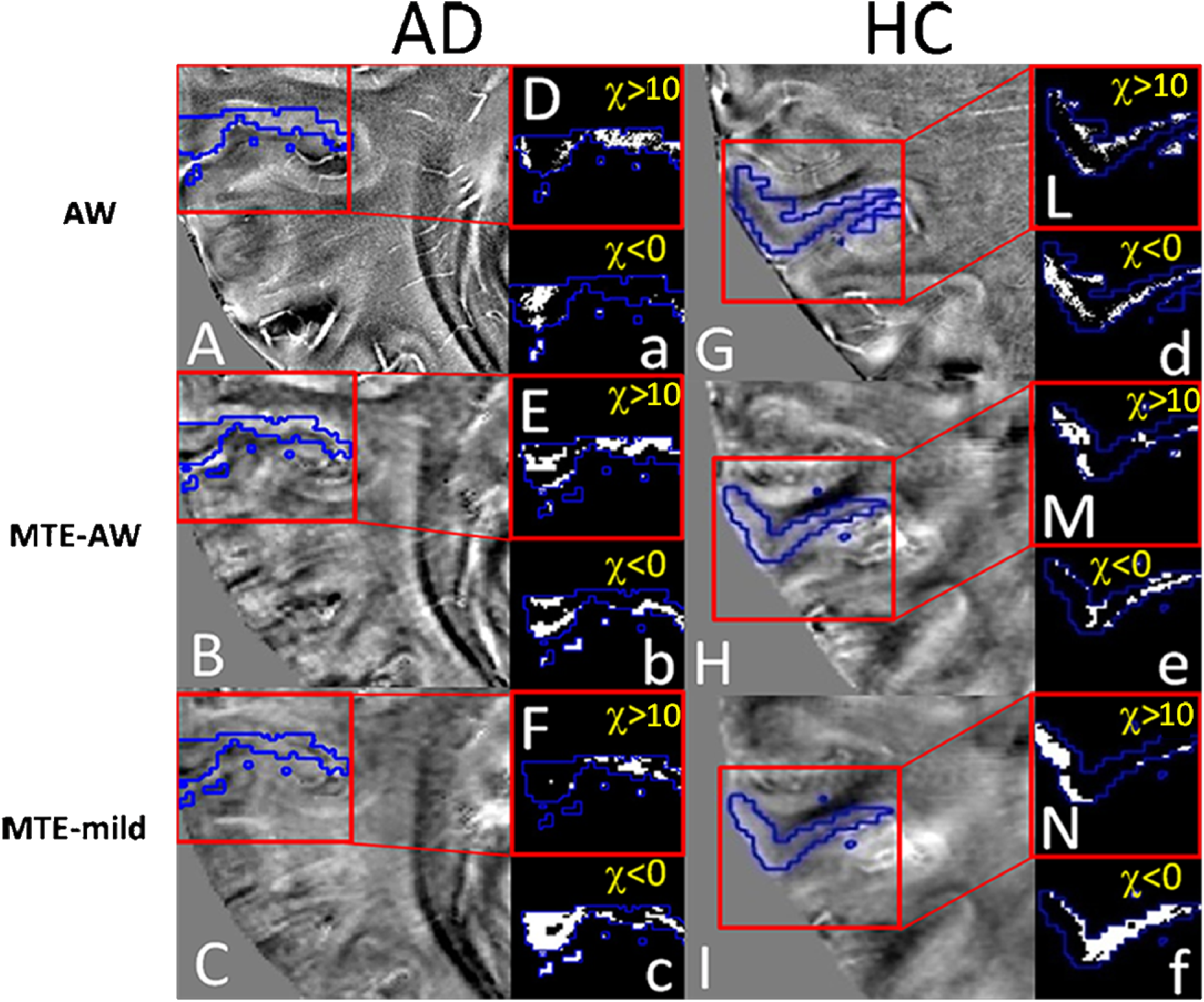
Paramagnetic and diamagnetic spatial patterns inside a representative ROI (*Supramarginal Gyrus posterior division,* blue) of an AD patient (D-F, a-c) and a HC (L-N, d-f), respectively obtained for χ>10 ppb and χ<0 ppb. “AW”, “MTE-AW” and “MTE-mild” refer to the maps obtained with the AW sequence, the MTE sequence with the AW-filter and with the mild-filter, respectively.

**Fig. 3.**
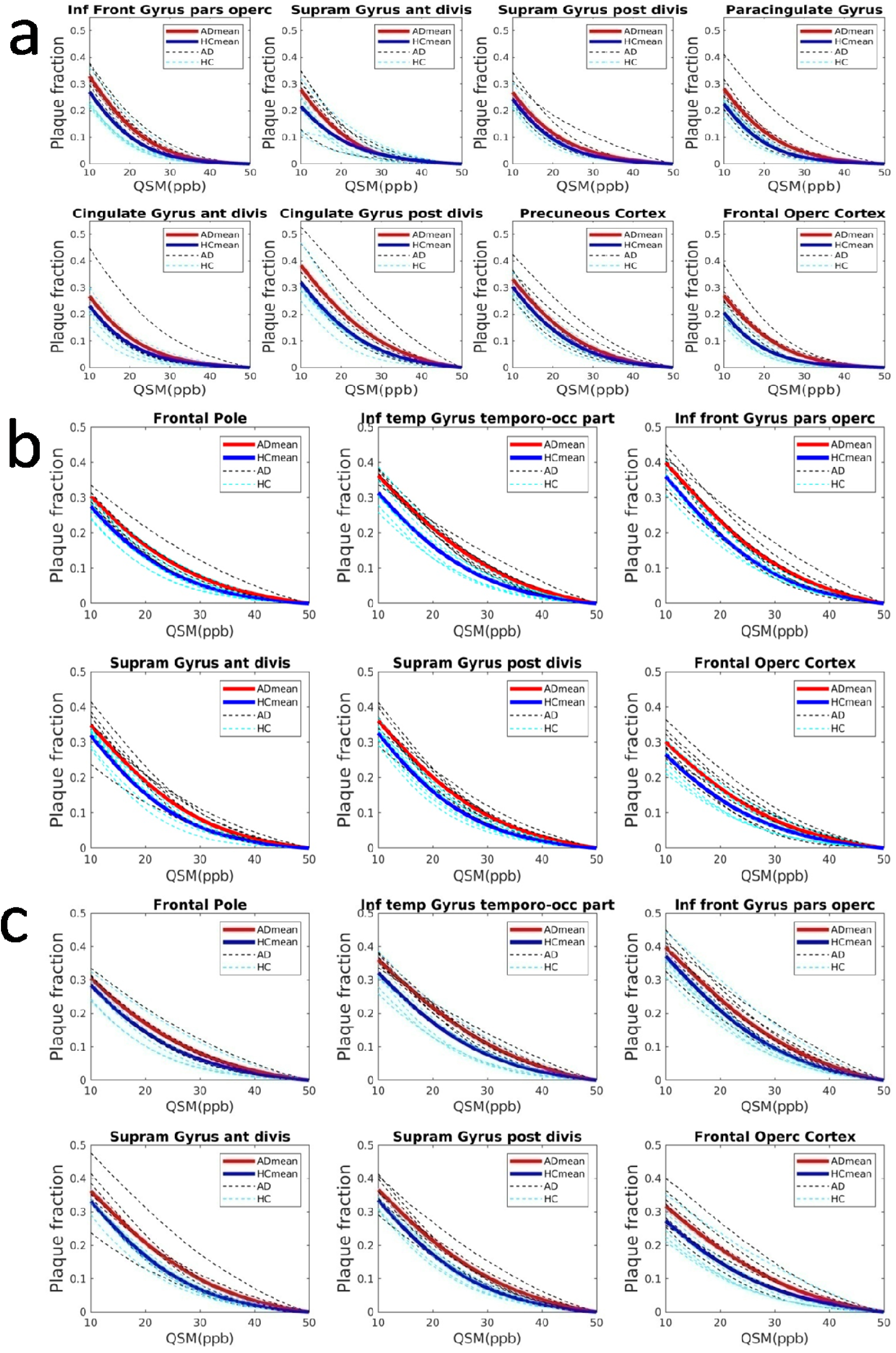
Mean paramagnetic load curves with a significant difference (p<0.05) between AD (red) and HC (blue) groups from ultra-high-resolution QSM (a), lower resolution QSM with the AW-filter (b) and with the mild-filter (c); black and cyan curves refer to single AD patients and controls, respectively. All methods enabled detection of cortical regions known to be involved in AD progression. Ultra-high resolution maps enabled detection of regions established as earliest Aβ accumulation areas, which were not detected at lower resolution. **Abbreviations: Inf Front Gyrus pars operc,** Inferior Frontal Gyrus pars opercularis; **Supram Gyrus ant divis,** Supramarginal Gyrus anterior division; **Supram Gyrus post divis,** Supramarginal Gyrus posterior division; **Cingulate Gyrus ant divis,** Cingulate Gyrus anterior division; **Cingulate Gyrus post divis,** Cingulate Gyrus posterior division; **Frontal Operc Cortex,** Frontal Operculum Cortex; **Inf temp Gyrus temporo-occ part,** Inferior Temporal Gyrus temporo-occipital part; **Inf front Gyrus pars operc,** Inferior Frontal Gyrus pars opercularis.

**Fig. 4.**
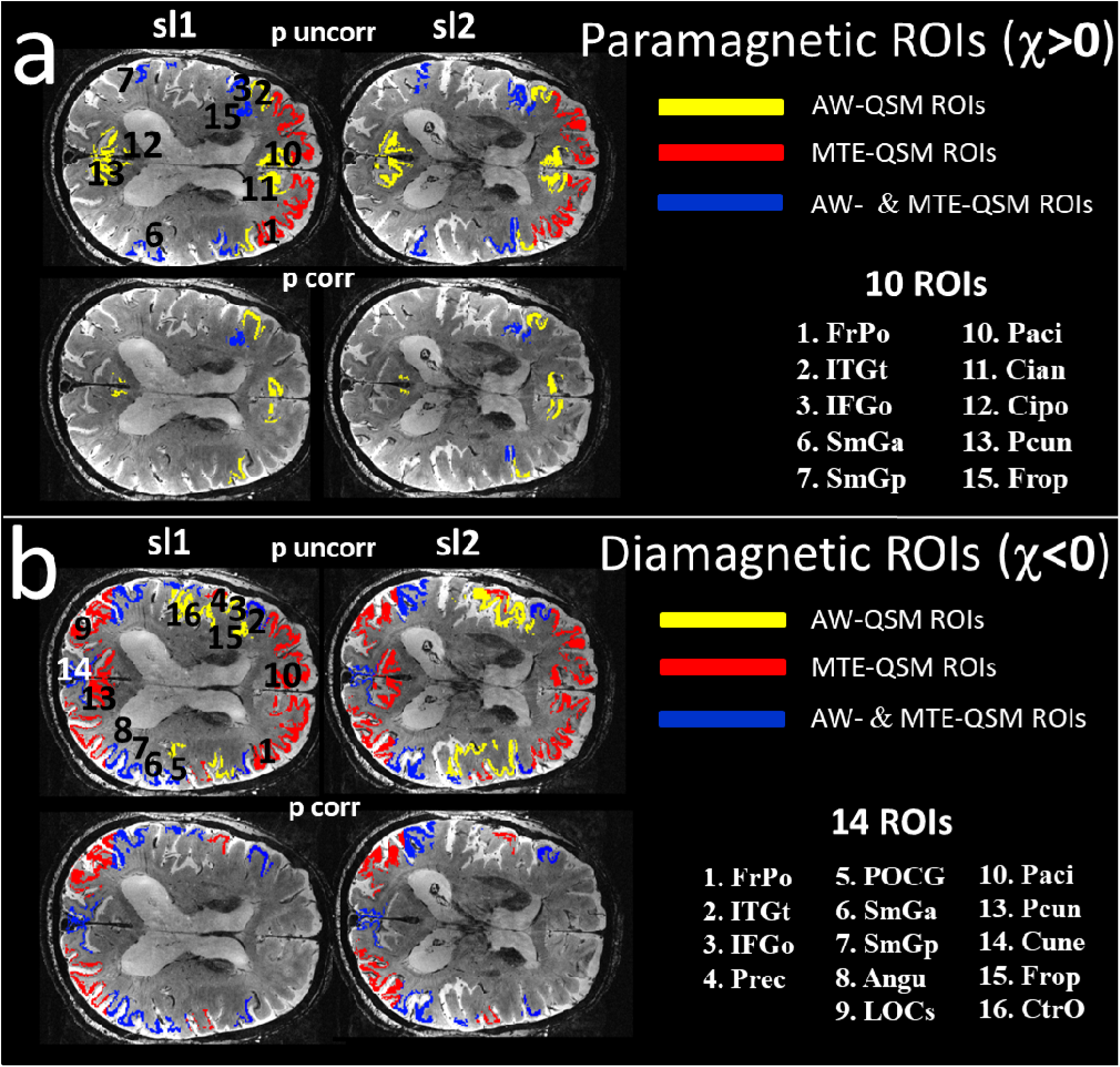
ROIs showing a significant difference between AD patients and HC groups based on paramagnetic load (a) and diamagnetic effects (b). **a)** Paramagnetic load detection in two different slices (sl1 and sl2) without Bonferroni correction (upper row) and with Bonferroni correction (bottom row). Red and yellow refer to the regions detected by only the AW technique and only the MTE maps, respectively. Blue refers to the ROIs whic were detected by both AW- and MTE- QSM regardless of the QSM effects (paramagnetic or diamagnetic). ROIs are overlaid on the magnitude image from the single-echo GRE of an AD patient. **1.** Frontal Pole. **2.** Inferior Temporal Gyrus temporo-occipital part. **3.** Inferior Frontal Gyrus pars opercularis. **6.** Supramarginal Gyrus anterior division. **7.** Supramarginal Gyrus posterior division. **10.** Paracingulate Gyrus. **11.** Cingulate Gyrus anterior division. **12.** Cingulate Gyrus posterior division. **13.** Precuneus. **15.** Frontal Operculum Cortex. **b)** Diamagnetic load detection in two different slices (sl1 and sl2) (upper row); after Bonferroni correction (bottom row). **1.** Frontal Pole. **2.** Inferior Temporal Gyrus temporo-occipital part. **3.** Inferior Frontal Gyrus pars opercularis. **4.** Precentral Gyrus. **5.** Postcentral Gyrus. **6.** Supramarginal Gyrus anterior division. **7.** Supramarginal Gyrus posterior division. **8.** Angular Gyrus. **9.** Lateral Occipital Cortex superior division. **10.** Paracingulate Gyrus. **13.** Precuneus. **14.** Cuneal Cortex. **15.** Frontal Operculum Cortex. **16.** Central Operculum Cortex. Please, refer to the Supplementary Information to visualize all 16 ROIs (Fig. S1).

**Fig. 5.**
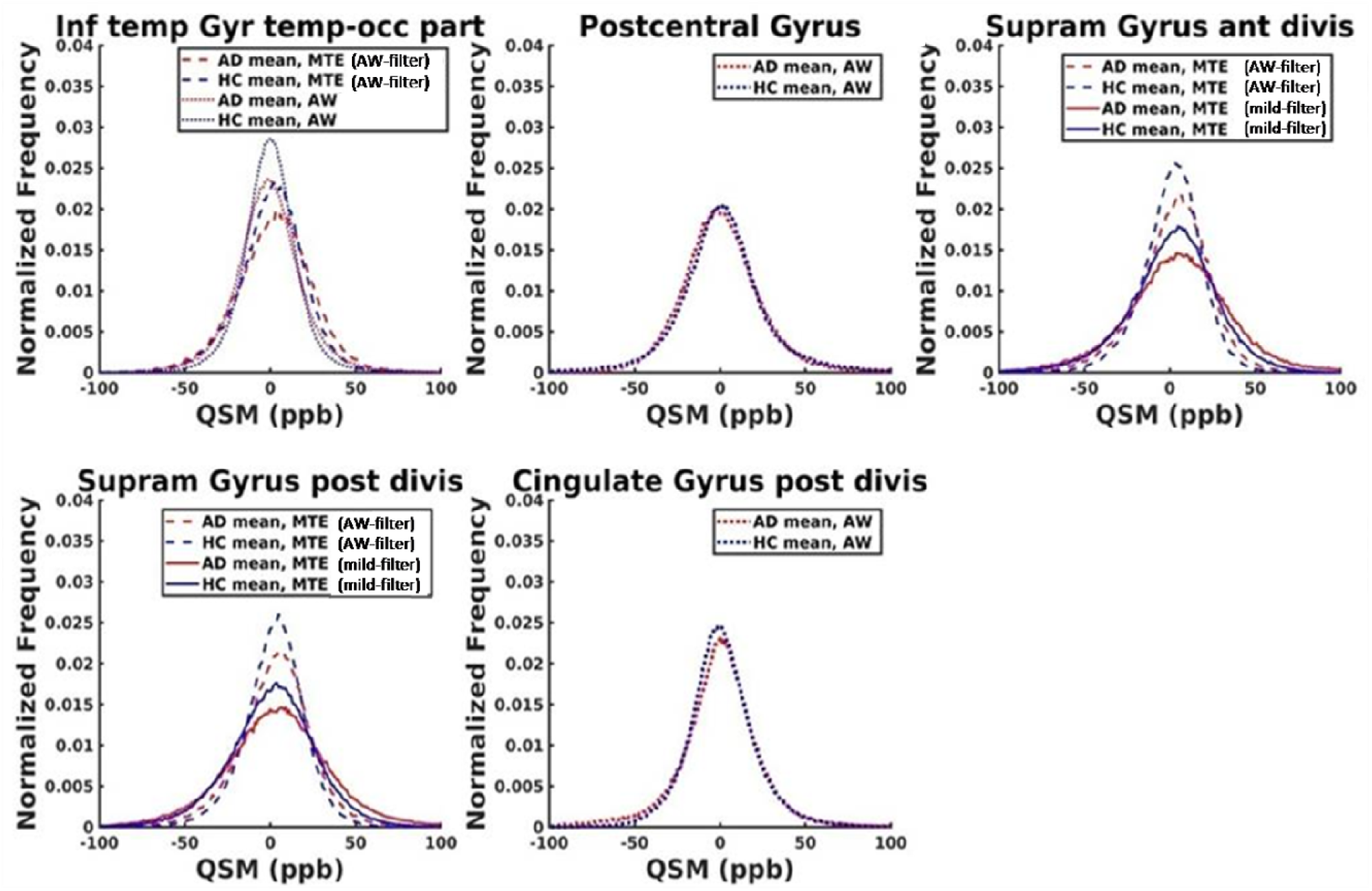
Histogram curves of QSM values showing a significant difference (p<0.05) between AD patients (red) and HC (blue) groups, obtained from ultra-high (dotted line) and lower resolution QSM, using the AW-filter (dashed line) and the mild-filter (solid line). **Abbreviations: ppb,** part per billion; **AW,** ultra-high resolution sequence; **MTE (AW-filter)**, MTE sequence with a AW-filter; **MTE (mild-filter),** MTE sequence with a mild-filter; **Inf temp Gyr temp-occ part,** Inferior Temporal Gyrus temporo-occipital part; **Supram Gyrus ant divis,** Supramarginal Gyrus anterior division; **Supram Gyrus post divis,** Supramarginal Gyrus posterior division; **Cingulate Gyrus post divis,** Cingulate Gyrus posterior division.

**Table 1.** ROIs showing a significant difference [corrected (p<0.05/32, (*)) and uncorrected (p<0.05) for multiple comparisons] between AD and HC based on para- (χ>0) and diamagnetic (χ<0) effects using the three methods.

| **16 Harvard**<br>**Cortical**<br>**ROIs** | **PARAMAGNETIC**<br>**DIFFERENCES**<br>**($\chi > 0$)**<br>**p-value** |  |  | **DIAMAGNETIC**<br>**DIFFERENCES**<br>**($\chi < 0$)**<br>**p-value** |  |  |
| --- | --- | --- | --- | --- | --- | --- |
|  | **AW-QSM**<br>**"AW-filter"** | **MTE-QSM**<br>**"AW-filter"** | **MTE-QSM**<br>**"Mild-filter"** | **AW-QSM**<br>**"AW-filter"** | **MTE-QSM**<br>**"AW-filter"** | **MTE-QSM**<br>**"Mild-filter"** |
| *Frontal Pole* |  | 0.00548 | 0.029 |  | 0.0019 | 0.0048 |
| *Inferior*<br>*Temporal*<br>*Gyrus temporo-*<br>*occipital part* |  | *0.00023 | *0.0012 | *0.0003 |  | 0.035 |
| *Inferior Frontal*<br>*Gyrus pars*<br>*opercularis* | 0.0028 | 0.008 | 0.022 | 0.026 |  |  |
| *Precentral*<br>*Gyrus* |  |  |  |  | 0.015 | *0.0006 |
| *Postcentral*<br>*Gyrus* |  |  |  | *3.3e-09 | 0.02 | *8.2e-05 |
| *Supramarginal*<br>*Gyrus anterior*<br>*division* | 0.028 | 0.014 | 0.0038 | *4.5e-15 | *2e-07 | *1.2e-06 |
| *Supramarginal*<br>*Gyrus posterior*<br>*division* | 0.02 | 0.004 | 0.0027 | *0.0001 | *4.7e-05 | *0.00014 |
| *Angular Gyrus* |  |  |  | *1.3e-05 | *1.7e-07 | *3.9e-09 |
| *Lateral*<br>*Occipital*<br>*Cortex superior*<br>*division* |  |  |  |  | 0.002 | *9e-05 |
| *Paracingulate Gyrus* | *9.5e-05 |  |  |  | 0.022 | 0.0032 |
| *Cingulate Gyrus anterior division* | 0.014 |  |  |  |  |  |
| *Cingulate Gyrus posterior division* | *0.0002 |  |  |  |  |  |
| *Precuneous Cortex* | 0.033 |  |  |  |  | 0.048 |
| *Cuneal Cortex* |  |  |  | *2.3e-08 |  | 0.037 |
| *Frontal Opercular Cortex* | *4.4e-06 | 0.008 | *0.0009 | 0.011 |  |  |
| *Central Opercular Cortex* |  |  |  |  | 0.044 | 0.021 |

**Abbreviations:** $\chi$ , susceptibility; **QSM**, quantitative susceptibility mapping; **AW**, acquisition-weighted sequence; **ROI**, region of interest; **AW-QSM**, susceptibility map obtained using the AW sequence; **MTE-QSM**, susceptibility map obtained using the MTE sequence.
**Notes:** Asterisks refer to regions detected after Bonferroni correction ( $p < 0.05/32$ ); $\chi > 0$ : paramagnetic load; $\chi < 0$ : diamagnetic effects;

Table 1 shows the ROIs with a consistent significant difference [corrected (p<0.05/32) and uncorrected (p<0.05) for multiple comparisons] between AD and HC detected using the three methods based on para- (χ>0) and diamagnetic (χ<0) effects. To correct for multiple comparisons we divided p=0.05 by 32, taking into account the number of common ROIs (16) and the considered magnetic effects (para- and diamagnetic). P values obtained after multiple comparisons correction are marked with an asterisk.

## 4. DISCUSSION

In this study, we explored paramagnetic and diamagnetic features of Alzheimer’s cortical microstructure that can be detected using in vivo QSM at 9.4T. We built upon and extended our previous results obtained in vivo at 9.4T and ex vivo at 9.4 and 14.1T [24]. In this way, we could explore QSM sensitivity in different settings, going from ultra-high resolution (37 µm isotropic voxel-size) achieved ex vivo at 14.1T throughout ultra-high resolution achieved in vivo at 9.4T (132x132x610 µm^3^) to the lower resolution used in the present in vivo study (375x375x800 µm^3^) at 9.4T. Specifically, in our previous work we observed a close link between QSM contrasts in ultra-high resolution maps and the spatial patterns in Anti-Aβ Immunohistochemistry of the same specimens. Based on that, we proposed a method to quantify cortical paramagnetic and diamagnetic effects to investigate the possibility of distinguishing AD from HC through the magnetic properties of the pathological tissue affected by β-Amyloid plaques using in vivo ultra-high resolution QSM at 9.4T.

In the present study, we confirmed the presence of different QSM contrasts across the cortex of a larger but still modest cohort of 7 AD patients and 7 controls at 9.4T. We also verified the existence of a consistent significant difference in the cortical paramagnetic and diamagnetic changes between AD and HC groups using the same ultra-high resolution AW approach as in the previous study. The long scan times required for the AW sequence allow for limited slab coverage of the brain (1.28 cm) that hampers the spatial registration to the whole brain anatomical reference image and limits the number of ROIs that can be included in the measurement. Therefore, the second aim of the present study was to explore the possibility of using the same method at a coarser spatial resolution with a 7.5-fold increase in axial coverage and a 30% reduction in scan time (MTE). This stepwise approach from ultra-high resolution ex vivo imaging at 14.1T to in vivo MRI at 9.4T consents exploring choices that may facilitate the clinical translation of such a QSM-based method for detection of β- Amyloid effects and verifying that this is still possible when using lower image resolutions and faster acquisition techniques.

### 4.1. Methodology and overall findings

In the present study, the ultra-high resolution AW approach [24] was used as the reference technique. However, the small extension of the AW-FOV allowed the inclusion of a limited number of ROIs (16 out of the 48 cortical regions available in the Harvard-Oxford brain atlas) that could be used for the analyses. For this reason, before acquiring the AW images, the FOV was carefully placed in such a way that the frontal, parietal, temporal and occipital cortices, corresponding to early Aβ accumulation areas [8, 9, 31], could be included. Results from the MTE maps were obtained in the same ROIs using two different background removal methods: a “strong” filter optimized for detection of fine details (“AW-filter”) as used for the AW approach [24] and a “mild-filter” widely used in the literature and known to restore the expected QSM contrast based on age-related estimates of the tissue iron-concentration [33].

We observed a distinct cortical pattern in AD patients compared to HC using ultra-high resolution AW-QSM in line with our previous results [24]. This pattern was confirmed at lower spatial resolution with both the AW- and the mild-filter. However, the AW-filter yielded a sharper and more localized pattern (with a slight loss in the spatial information at lower resolution) as well as a greater GM/WM contrast compared to the mild-filter, which instead showed a substantial loss of local field information across the cortical pattern of all subjects. On the other hand, the mild-filter exhibited an enhanced QSM contrast in the subcortical nuclei, in line with literature [19–23], which was less intense with the AW-filter and almost lost at higher resolution. The increased QSM contrast obtained with the mild-filter is also consistent with its improved iron-dependent contrast recently observed in healthy subjects [33] and is relevant for measurements of the basal ganglia in AD [18]. In the cortex, where Aβ protein start to aggregate [8, 9, 31], higher resolution maps enabled the detection of eight out of sixteen common regions with a significant difference between AD and HC groups based on the paramagnetic load. Four of these regions were also detected by the MTE maps, while two additional regions were only detected with the lower resolution data. As to the diamagnetic effects, six ROIs were detected regardless of the spatial resolution while six more regions were only detected with the MTE maps and other two only by the AW maps. Overall, the diamagnetic changes were more prominent than the paramagnetic effects, regardless of the spatial resolution. QSM histograms of the two groups obtained from both the MTE and the AW sequences were significantly different (p<0.05) in five out of sixteen ROIs based on a two-sample Kolmogorov-Smirnov test.

### 4.2. Method-dependent differences in QSM

The spatial distribution of the magnetic effects (paramagnetic and diamagnetic) observed in the susceptibility maps presented some differences depending on the voxel-size and the filter used. Specifically, the paramagnetic effects observed in the highly detailed ultra-high resolution maps (AW maps) showed a spatial pattern with small and localized sources likely reflecting iron bound to the β-Amyloid plaques. Similar patterns were also observed for the diamagnetic signal changes at this spatial resolution. On the other hand, the magnetic effects revealed in the MTE maps appeared smoothed and larger with slightly different spatial patterns compared to the AW approach. This may be caused by a joint action of the inevitable spatial under sampling of the local susceptibility sources within the tissue microstructure and partial volume effects that are manifest at lower resolution. Furthermore, while the AW maps showed an unambiguous contrast between diamagnetic and paramagnetic sources inside the segmented regions these details were not visible at lower resolution where, in contrast, a smoothed paramagnetic signal was dominant. Taken together, the under sampling effects and the loss of local diamagnetic information at lower resolution likely contributed to the slightly different spatial distribution of the effects observed in the MTE maps compared to the AW maps, using the same filter. This difference was further enhanced using the mild filter with its stronger QSM contrast. Nevertheless, despite the ‘smoothing’ effect observed at a lower resolution, the presence of paramagnetic sources could still be revealed. Indeed, as we previously investigated through a simple biophysical model based on histology [42], in principle, the global field effects caused by local iron-rich deposits can be revealed up to 1 mm voxel-size, although, in this case, only the combined effects of several sources (plaques) can be detected. These observations suggest that the spatial patterns observed in the higher resolution maps (AW maps) may reflect the local sources of the detected signal changes while the effects revealed with the other techniques (MTE maps) likely resembled combined effects of several plaques with possible partial volume effects. Another possible reason for the slightly different patterns of the magnetic effects observed in the AW and the MTE maps is the existence of artifacts in areas with strong field in-homogeneities, such as the frontal areas and subcortical regions, which particularly affected the MTE sequences compared to the ultra-high resolution maps, where these artifacts were either less enhanced or not observed at all. To minimize such issues, we used a navigator echo approach to correct for field fluctuations during 3D- GRE acquisition. However, unlike results obtained with R_2_* mapping [32] the quality of our phase-contrast-based quantitative maps did not substantially improve with the navigator approach. This may have contributed to the detection of signal changes in the MTE-QSM which were not detected by the AW technique. For this reason, we should only rely on those regions that were also detected by the reference AW technique. The histogram analysis of the MTE maps always provided broader curves and lower peaks compared to the AW-based maps as well as greater absolute susceptibility values in patients compared to HC. This effect was more pronounced using the mild filter. In addition, we observed a consistent slight paramagnetic shift towards high QSM values in the histogram curves with higher values in AD patients than HC. Also in this case, this could be explained by the over-smoothed signal obtained using a too low regularization (mild-filter) at a coarser resolution.

### 4.3. Confirmation of previous AWI-based study and reliability of MTE method

Taken together, these observations concerning the paramagnetic load, on the one hand, and the diamagnetic effects, on the other hand, are in line with our previous ex vivo results [24]. These showed different QSM contrasts across the cortical depth (in a frontal area of the cortex) that distinguished AD from HC consisting of two main bands (a broad paramagnetic band and a narrow strongly diamagnetic band) in AD, and three main bands (two large diamagnetic bands surrounding a central narrow paramagnetic band) in HC. According to that, we expected to detect distinct profiles for the different magnetic properties across the cortex of the two groups. Indeed, since QSM can distinguish between paramagnetic and diamagnetic sources [43] we aimed to detect iron bound to the plaques (paramagnetic), on the one hand, and cortical microstructural alteration of myelin (diamagnetic) as well as other possible mechanisms involved in the Aβ (e.g. diamagnetic elements of the plaques) and, possibly, tau-protein accumulation, on the other hand. Regardless of the method used, we observed four ROIs with significant differences between AD and HC based on the paramagnetic load, and six ROIs based on the diamagnetic effects. The ability to detect similar changes in some regions using both the AW and the MTE-QSM independently of the magnetic effects (para- and diamagnetic) suggests that results obtained with the validated technique (AW approach) do translate to a coarser resolution, and we can rely on those regions. Furthermore, all the regions showing significantly higher paramagnetic values in AD than HC, regardless of the image resolution, have been reported as early Aβ accumulation areas according to a recent study on the connection between PiB SUV ratio and Thal amyloid phases across the Alzheimer’s disease progression [44].

On the other hand, the AW technique enabled the detection of four additional regions with higher paramagnetic values in AD than HC which were not captured at lower resolution. These included two of the earliest Aβ accumulation areas (precuneus and posterior cingulate cortex (PCC)), based on recent studies combining Aβ-PET imaging with CSF test [13, 45]. This observation suggests that the AW technique may be particularly advantageous for the early detection of Aβ based on paramagnetic effects. Differences in QSM sensitivity for detection of paramagnetic effects may depend on the specific iron level which might be lower in the cortex than subcortical areas, as recently reported in a large clinical cohort of AD patients [23] using lower resolution QSM (0.52 × 0.52 mm^2^). On the other hand, regions that were only detected by the MTE sequence and not by the AW technique, regardless of the magnetic effects (para- and diamagnetic), may have been hampered by residual artifacts that manifested despite the use of a navigator echo approach to correct B_0_ fluctuations in 3D-GRE sequences.

### 4.4. Contributions from veins

The contribution of veins may hamper the detection of more local QSM effects. An accurate estimation of the vein-like confounders to be excluded can be achieved using a combined contrast of SWI and QSM [46]. However, with this approach, there is a risk to include non-local extra-vascular effects coming from voxels close to vessels orthogonally oriented to the field that can contribute to the SWI images. This question is highly important and deserves to be studied in the future. In the present study, we employed the same QSM-based method as in our previous study. It relies on the observation that paramagnetic effects greater than 50 ppb in in vivo human brain likely reflect vein contributions [27]. Therefore, in a first step, we excluded this contribution to discard sources likely not related to the Aβ plaques. The remaining susceptibility values (up to 50 ppb), which provided a good match between the spatial pattern of ex vivo QSM paramagnetic effects and the Aβ plaques distribution in the histological sections as a function of QSM cutoffs (from 10 to 50 ppb), were therefore included in the analyses. A possible dependence of the vein contribution on the category (AD or HC) was then explored by computing and comparing the vein fractions of the two groups for each method but was found to be non-significant.

### 4.5. Comparison with previous studies

The use of QSM for discerning AD from HC has been recently proven at 7 and 3T based on QSM changes observed in the subcortical nuclei [19–23]. With similar approaches QSM enabled the distinction of amnestic MCI from HC by detecting altered QSM values in the precuneus and allocortex regions at 3T [47], thus showing that QSM is an emerging valuable tool for the clinical use. Our QSM-based method always detected cortical regions consistent with Aβ accumulation areas as reported in several recent studies based on Aβ PET and Aβ CSF, like the *posterior* and *anterior cingulate cortex*, the *precuneus*, the *supramarginal gyrus*, the *inferior*, *middle* and *superior temporal lobes*, the *pars opercularis*, the *frontal pole* and *superior frontal lobe* [45, 48–53]. Other studies reported a decrease in the stiffness of the *precuneus*, the *operculum* and the *precentral gyrus*, as well as the *middle* and *superior temporal gyri*, using voxel-based MRE (Magnetic resonance elastography) in AD patients [54]. Also, it has been proven that atrophy and synapse loss occurs after Aβ accumulation in AD [55] and that atrophy in the *frontal pole* is associated with aberrant motor behavior and, in addition, a decreased volume of the *inferior frontal gyrus* is associated with behavioral symptoms (like disinhibition and elation) in Mild Cognitive Impairment, Alzheimer’s Disease, and Frontotemporal Dementia [56]. The *opercular frontal regions* are involved in the atrophy pattern of the AD-related logopenic variant of Primary Progressive Aphasia (lvPPA) [57]. These findings give support to our observations, since all the regions we found to enable distinguishing AD patients from HC based on estimated Aβ plaque-load are known to be involved in AD progression. Other groups reported the *precuneus* and the *posterior cingulate* cortex as the regions with the highest starting level of Aβ accumulation (up to 20 years before the estimated onset of AD) and with the fastest initial increase over the estimated time of the disease [58], succeeded by multiple *frontal lobe* regions [59].

### 4.6. Limitations of the present work

Our work has some limitations. Although our previous results were confirmed [24], the sample size was still very small and we could not generalize our findings to a larger population of subjects. This might also have diminished the power of our method in discerning the two groups. The lack of PET imaging for validating our results was another limitation of this study, especially considering the small size of our sample. In addition, healthy subjects did not undergo the CSF test to exclude amyloidal pathology or a prodromal stage (MCI). However, if the healthy subjects had Amyloid pathology or were in the prodromal stage, they would have necessarily had plaque-related signal alterations which would have been detected by our method. This would have significantly diminished the differences between HC and AD, since Aβ accumulation develops along a sigmoid curve with a very fast increase in the preclinical stage (HC), a very slow increase in the prodromal stage (MCI) and a plateau in the final stage (dementia). In contrast, we observed considerable differences in the QSM effects between the two groups. Another limitation of our study was the long scan time required for the AW technique which allowed for limited slab coverage of the brain. This restricted the number of ROIs that could be included for the analyses. Therefore, a special care was taken to place the slab to include representative ROIs. In addition, the limited slab thickness of the AW-MRI data hampered the direct registration with the whole-brain anatomical reference image. Therefore, the spatial transformation of the Atlas-based regions from the MP2RAGE space to the ultra-high resolution AW-QSM required a multi-step processing. Finally, for safety reasons, many subjects and patients who were willing to participate in the study had to be discarded since they had metallic material in their teeth, or joint implants.

### 4.7. Future outlook for clinical translation

The clinical translation of a QSM-based method for the examination of cortical regions would benefit from adaptations of the measurement techniques to the clinical setting. Our results suggest that the acquisition of single-echo data [40] rather than multiple echoes as well as the choice of a few representative ROIs instead of the whole brain may suffice to distinguish AD from HC based on the proposed QSM method. Such choices can significantly reduce the scanning times and may eventually be traded with an increased spatial resolution at lower field strengths of 7T and, possibly, at 3T. In this respect, it should be emphasized that that the diamagnetic changes were in general more prominent than the paramagnetic effects regardless of the spatial resolution, and their detection was more straightforward than paramagnetic changes. Therefore, the diamagnetic effects, possibly driven by Aβ as well as tau-pathology accumulation as suggested in an AD mouse model [17], may prove to be of particular interest at lower magnetic field strengths and lower resolutions currently available in the clinic.

## 5. CONCLUSIONS

With this work, we explored the cortical features of Alzheimer’s brain that can be detected in vivo at

9.4T using QSM at two spatial resolutions. The proposed QSM-based method can detect signal changes likely related to iron bound to Aβ plaques, on the one hand, and to other mechanisms involved in the Aβ and, possibly, tau-protein accumulation, on the other hand. This study points toward developing more accessible clinical methods to non-invasively detect effects of Aβ accumulation in AD patients.

## Data Availability

All data produced in the present study are available upon reasonable request to the authors.

https://github.com/ElisaTuzzi/9.4TQSM_Alzheimer

## ACKNOWLEDGMENTS

We gratefully acknowledge the EU-LAC Foundation [Grant #16/T01-0118] and the German Research Foundation Max Planck Society ERC for financially supporting this study. The funders were not involved in data analysis or interpretation of the results.

